# Reconstructing the range-normalized terminal QRS from V1 and V6 using the time-embedded 3D ECG

**DOI:** 10.64898/2026.11.14.26368083

**Authors:** Alejandro Jesús Bermejo Valdés, Jessica Archer Jiménez

## Abstract

**Objective:** Can a fixed V1/V6 projection of the time-embedded 3D ECG estimate the final 40 ms of V2–V5 more accurately than a fitted single lead?

**Methods:** We used simultaneous recordings from the Lobachevsky University Electrocardiography Database (LUDB), with cardiologist-annotated QRS complexes in all six chest leads. We fitted both models in 118 patients and tested them in 80 others. Root mean squared error (RMSE) is expressed in percentage points (pp) of the target lead’s 10-second amplitude range.

**Results:** In 733 test beats, patient-weighted terminal RMSE fell from 12.58 to 10.64 pp (difference 1.94 pp, 95% bootstrap confidence interval 1.30 to 2.55). Reductions were 3.23, 2.85 and 1.35 pp in V3, V4 and V5. In V2 the difference was 0.35 pp (95% interval *−* 0.35 to 1.05). In 702 matched beats from 77 patients, the overall advantage was 2.22 pp at the automatic V1/V6 end and 1.92 pp at the annotated target end. Of 1,498 beat–lead pairs with whole-QRS correlation *≥* 0.95, 28.84% still had terminal RMSE of at least 10 pp.

**Conclusions:** A fixed V1/V6 projection improved offline estimates of relative terminal amplitude in V3–V5 without fitting the target lead in the new patient. The gain was largest in V3 and V4 and uncertain in V2. Whether a reconstructed trace preserves a clinically recognizable notch or slur remains untested.

## Introduction

When a reduced-lead recording includes V1 and V6 but omits the intervening chest leads, the clinician sees the terminal QRS in two precordial leads and must infer how it might appear in the others [1]. For example, when evaluating an early repolarization pattern in V4 or V5, a reader examines the final downslope of the R wave for a notch or slur [2]. A reconstruction may follow the dominant R wave closely and still miss a smaller, later deflection. This makes the final 40 ms a clinically relevant segment in which to test a reconstruction.

The time-embedded three-dimensional ECG plots V1 and V6 against time. Changing the projection angle changes the contribution of each lead to the resulting trace. Earlier work used this representation to examine precordial waveforms in STAFF III recordings [3], R-wave progression across viewing directions [4] and apparent J waves in V6 [5]. Here the angle has a different use. If an angle learned for V4 in one group can estimate the terminal V4 waveform in another, the same two recorded signals can provide an estimate of a lead unavailable to the reader, without individual calibration of the angle. Nelwan et al. studied reduced lead sets, and Horáček et al. tested transformations combining limb electrodes with pairs of chest electrodes [1, 6]. We asked a narrower question about the two precordial inputs alone. For which targets does the fixed projection reduce error over the final 40 ms compared with a fitted single lead? Does the comparison change when the end of the scoring window is set by V1 and V6?

## Methods

### Recordings and participants

LUDB version 1.0.1 contains a 10-second, simultaneous 12-lead ECG sampled at 500 Hz for each of 200 participants. It includes healthy volunteers and patients with cardiovascular disease. Two LUDB cardiologists examined each lead visually and agreed on the wave onsets, peaks and ends [7, 8]. We assigned 120 participants to development and 80 to testing by sorting a hash of the public record identifier before refitting either model (Supplement, Section S1). All rhythms were eligible. Each participant contributed to one partition. We used the public waveforms and annotations without contacting participants. The LUDB investigators report written informed consent for the original data collection and approval by the Lobachevsky University institutional review board (No. 23, 19 October 2017) [8].

### Beat confirmation and terminal window

For each annotated V3 QRS peak, we required a unique matching peak within 100 ms in each of the other five precordial leads and a unique match back to V3. The six annotated QRS intervals had to overlap. For every target lead, all 20 terminal samples had to lie inside its own annotated QRS. The criterion establishes the identity of a common beat while allowing depolarization to appear to end at different times across leads. The automatic detector played no part in selecting the primary cohort.

The release-specific scale is important here. In all 2,400 record–lead combinations, the WFDB gain in counts per mV equalled the digital maximum minus the minimum over that recording. Applying the header therefore gives an identical peak-to-peak range of 1 mV in every channel. We express errors in percentage points (pp) of each target’s 10-second range. One pp corresponds to 10 nominal *μ*V in the distributed files. An independent physical calibration is unavailable. This scale uses the entire recording, including each measured target for the reference. Section S10 documents the audit.

We rejected segments with missing samples, a digital rail or a flat precordial QRS (Section S1). If *o* is the earlier annotated V1 or V6 onset, each channel was centred on its own median over samples *o−* 25, …, *o−* 6. If *k*_*j*_ is the target’s closing annotation, its final 40 ms comprise the 20 samples *k*_*j*_ *−* 19, …, *k*_*j*_. Both models used unfiltered samples. The single-lead comparator used one channel for amplitude but shared the V1/V6 onset for centring. This centring interval can overlap an annotated P wave. In a test patient the target supplied the reference waveform and its boundaries. Only the two input waveforms entered the fixed predictions.

### Reconstruction models

Let *x*_1_(*t*) and *x*_6_(*t*) denote baseline-corrected, range-normalized inputs. The time-embedded trajectory is (*x*_1_(*t*), *x*_6_(*t*), *t*). For target *j*, the projection (A) and single-lead comparator (B) were

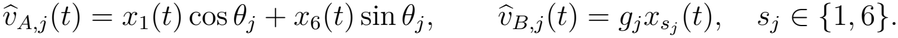

Weestimated one angle per target. For B, we fitted a signed gain for each candidate input and retained the input with the lower development error. Both intercepts were zero after baseline subtraction. The fitting criterion was mean squared error in normalized amplitude with equal weight for each participant, for each beat within participant and for each sample within a window. All choices of angle, input lead and gain were fixed before scoring the 80 test participants. Section S2 gives the coefficients.

### Outcomes and statistical analysis

For each beat and target, we calculated the square root of the mean of the 20 squared differences between predicted and measured range-normalized amplitude. We averaged these RMSEs across beats within participant and target, then across the four targets and finally across participants. The primary effect was the single-lead RMSE minus the projection RMSE. A positive value means that the projection was closer to the recorded waveform. We obtained percentile 95% confidence intervals (CIs) from 5,000 paired resamples of entire test participants, conditional on the fitted coefficients. In a subsequent analysis we tested the mean patient difference with a two-sided paired *t* test. Holm adjustment controlled the familywise error rate at 0.05 across the four lead-specific tests [9]. The CIs in Table 1 are marginal for each estimate.

**Table 1:** Terminal RMSE and paired differences in 80 test participants. Errors and differences are in pp. The global *p* value is unadjusted. Lead-specific *p* values are Holm-adjusted. A positive difference favours the projection.

| Target | Projection A | Single lead B | B minus A | 95% bootstrap CI | *p* |
|---|---|---|---|---|---|
| Overall | 10.64 | 12.58 | 1.94 | [1.30, 2.55] | 4.82 × 10<sup>−8</sup> |
| V2 | 10.68 | 11.03 | 0.35 | [-0.35, 1.05] | 0.330 |
| V3 | 12.78 | 16.01 | 3.23 | [2.12, 4.33] | 7.49 × 10<sup>−7</sup> |
| V4 | 11.64 | 14.49 | 2.85 | [1.83, 3.81] | 1.57 × 10<sup>−6</sup> |
| V5 | 7.46 | 8.81 | 1.35 | [0.76, 1.91] | 3.46 × 10<sup>−5</sup> |

For the timing comparison, a two-input detector located a candidate complex from 8–30 Hz signal energy and estimated the end from a sustained fall in slope activity (Section S3). We used the first qualifying low-slope sample as the exclusive automatic stop. The anchor had to lie inside all six annotated intervals, and all target leads had to pass the signal-quality checks. Each matched beat was scored twice, changing only the end of the 40-ms window. The baseline and coefficients remained fixed. We treated the four timing tests as a separate exploratory family. Other sensitivity analyses compared equal-gain restrictions, fitting directly to window RMSE, omission of individual patients, fivefold patient-level validation and a bootstrap with independent resampling of development and test participants. The within-window least-squares fit used the measured target and gives a retrospective bound.

### Analysis history

The restriction to six-lead cardiologist annotations followed an earlier analysis of automatic QRS timing. The first author, who introduced the time-embedded ECG framework, previously studied geometric descriptors in these same 200 LUDB recordings [10]. PTB-XL and Shaoxing–Ningbo had no role in the present annotated analysis. We recorded the revised eligibility rule, patient partition and primary endpoint before refitting. The paired tests, multiplicity adjustment and bootstrap with refitting were added after the effect estimates were known. The protocol and statistical addendum accompany the code.

## Results

### Population

Of 1,831 V3 QRS markers, 14 lacked a complete V3 onset–peak–end triplet. Of the remaining 1,817 complexes, 1,800 from 198 participants met the six-lead criteria. The remaining 17 had no complete corresponding annotation in at least one other lead. Two development participants had no eligible complex. Development therefore included 1,067 beats from 118 participants (median age 56.5 years, 49 women), and testing included 733 beats from 80 participants (median age 54.5 years, 35 women). Each eligible beat contributed four target windows. Counting all marked peaks, 20 of 77 in records labelled with ventricular pacing were ineligible, compared with 11 of 1,754 in the other records. These labels describe the recording as a whole.

### Reconstruction on annotated windows

Across the four target leads, mean terminal RMSE fell from 12.58 to 10.64 pp (Table 1). The difference of 1.94 [1.30, 2.55] pp represents 15.46% of the comparator’s mean error. 63 of 80 patients had a lower four-lead mean error with the projection. V3 showed the largest mean reduction, followed by V4 and V5. The small estimated reduction in V2 was compatible with either a gain or a loss. With both gains free, mean RMSE was 10.41 pp across the four leads (model C). Projection RMSE remained 12.78 pp in V3 and 11.64 pp in V4. Figure 1 illustrates the projection in a held-out patient.

**Figure 1:**
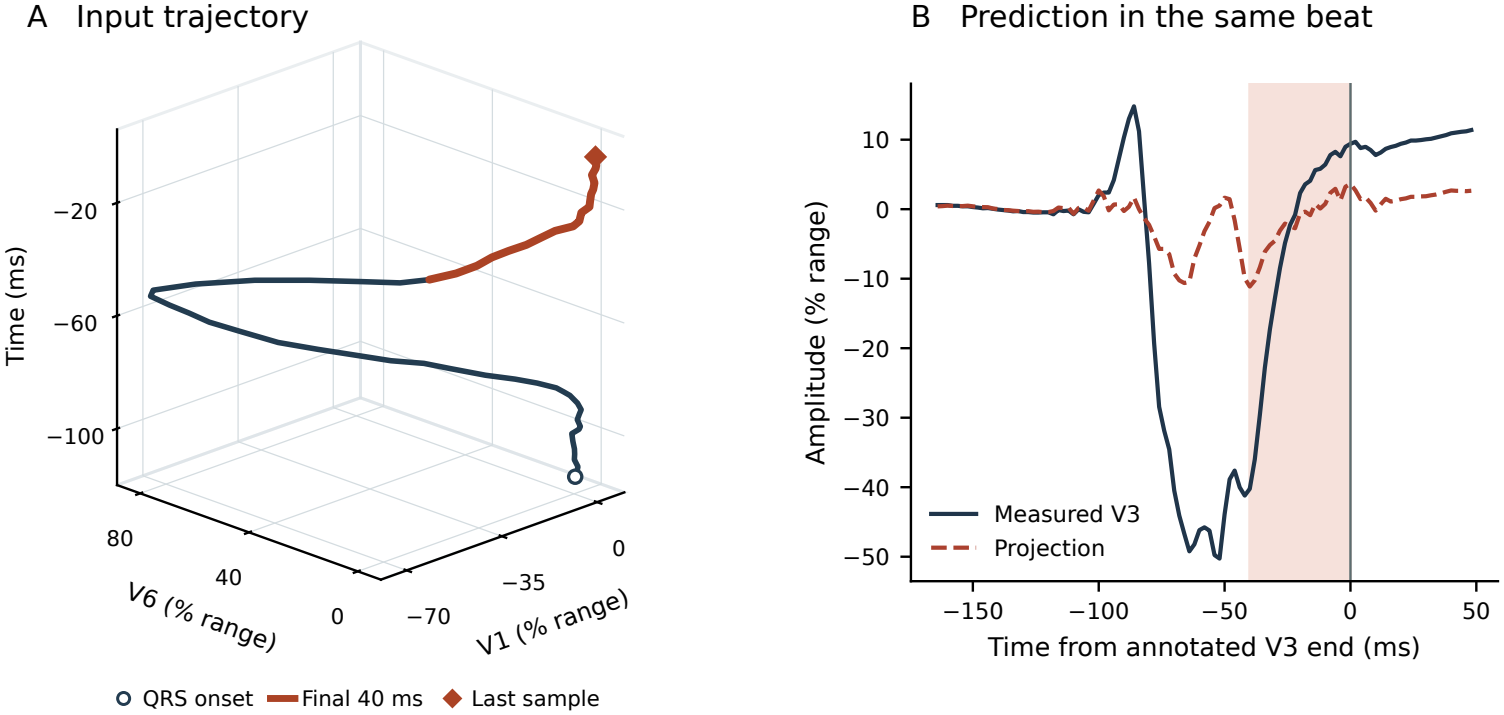
Time-embedded ECG and its fixed V3 projection. Panel A shows the range-normalized V1 and V6 signals throughout the annotated QRS of LUDB record 28 (V3 peak at sample 2236). The open circle marks the annotated V3 onset. The curve runs out and returns as time advances upwards. The orange segment contains the final 20 samples, ending at the diamond. Panel B shows the projection and the simultaneously measured V3. The shaded interval contains samples from *−*40 to *−*2 ms. Zero is the first sample after the annotated closing sample. This beat was nearest the median V3 test error.

### Sensitivity to the QRS end

Of 733 eligible annotated beats and 833 automatic detections with valid bounds and signals, 702 matched in 77 participants (2,808 beat–lead pairs). The automatic window extended beyond the local annotated QRS end in 831 pairs. In 43 pairs the two terminal windows shared no samples. Of 16 eligible beats in the three test recordings labelled with ventricular pacing, only five from one person contributed to this comparison, compared with 697 of 717 beats in the other recordings. The automatic end preceded the target-specific annotated end by 4.31 ms on average across the four leads. The mean absolute discrepancy was 8.10 ms. On average, both models had lower RMSE when the same beats were scored at the annotated end (Table 2). The resulting overall change in their relative advantage was −0.29 [−0.60, 0.01] pp (*p* = 0.065). V3 showed the largest estimated change, −1.07 pp (nominal *p* = 0.015, Holm-adjusted *p* = 0.060). In V2, the estimated advantage changed from −0.07 to 0.34 pp. The marginal interval at each endpoint included zero. Figures S1 and S2 show measured excursions that extend beyond the input-defined end.

**Table 2:** Effect of changing the end of the QRS in the same 702 beats. Each pair of entries gives the automatic value first and the annotated value second. Change is the annotated minus automatic advantage. Values and patient-bootstrap 95% intervals are in pp.

| Target | Projection A | Single lead B | Advantage (B minus A) | Change [95% CI] |
|---|---|---|---|---|
| Overall | 13.02 / 10.86 | 15.23 / 12.78 | 2.22 / 1.92 | -0.29 [-0.60, 0.01] |
| V2 | 12.60 / 10.87 | 12.53 / 11.20 | -0.07 / 0.34 | 0.41 [-0.06, 0.87] |
| V3 | 16.97 / 13.11 | 21.14 / 16.21 | 4.17 / 3.10 | -1.07 [-1.93, -0.24] |
| V4 | 14.43 / 11.88 | 17.78 / 14.78 | 3.34 / 2.90 | -0.44 [-0.96, 0.08] |
| V5 | 8.07 / 7.59 | 9.49 / 8.94 | 1.42 / 1.35 | -0.06 [-0.27, 0.15] |

### Sensitivity to fitting and influential observations

When each candidate single lead was centred on its own annotated onset and the comparator was refitted in development, the projection’s mean advantage was 1.99 pp. The primary baseline overlapped a target QRS in 107 of 733 test beats. Recentring before the earliest of all six marked onsets and refitting in development gave a mean reduction of 1.93 [1.29, 2.54] pp. Moving the input-based reference 20 ms earlier gave 1.89 [1.25, 2.51] pp (Section S11). Training both methods directly on mean window RMSE preserved the reductions in V3–V5. In V2 the interval continued to include zero with matched gain freedom and with the alternative loss (Table S2). In V2, imposing unit gain on both models gave *−* 0.09 pp (95% CI *−* 1.19 to 1.05). Free gains for both gave 0.37 pp (*−* 0.36 to 1.09). After excluding the patient with the highest mean projection error across the four leads, the V2 difference was 0.30 [−0.37, 0.98] pp. In a descriptive fivefold patient-level cross-validation the difference was 0.56 pp in V2 and 2.00 pp across the four leads (Table S3). When both cohorts were resampled independently and both models refitted in 4,000 replicates, the global 95% interval was [1.29, 2.63] pp. Omitting one test participant at a time gave overall estimates within [1.87, 2.06] pp. Excluding all three test recordings labelled with ventricular pacing gave a reduction of 1.89 [1.24, 2.52] pp in the remaining 77 participants (Section S6).

### Full-QRS correlation and terminal waveform error

A full-QRS Pearson correlation of at least 0.95 coexisted with a terminal projection RMSE of at least 10 pp in 432 of 2,932 beat–lead pairs (14.73%). Of the 1,498 pairs reaching that correlation, 28.84% had terminal RMSE at or above 10 pp. In V3, 70 of 159 high-correlation pairs (44.03%) still met the terminal error threshold. Table S5 gives results across thresholds and Table S8 gives the counts for each lead. Figure 2 shows the measured and reconstructed V3 waveforms at two percentiles of test error and for a beat meeting both thresholds.

**Figure 2:**
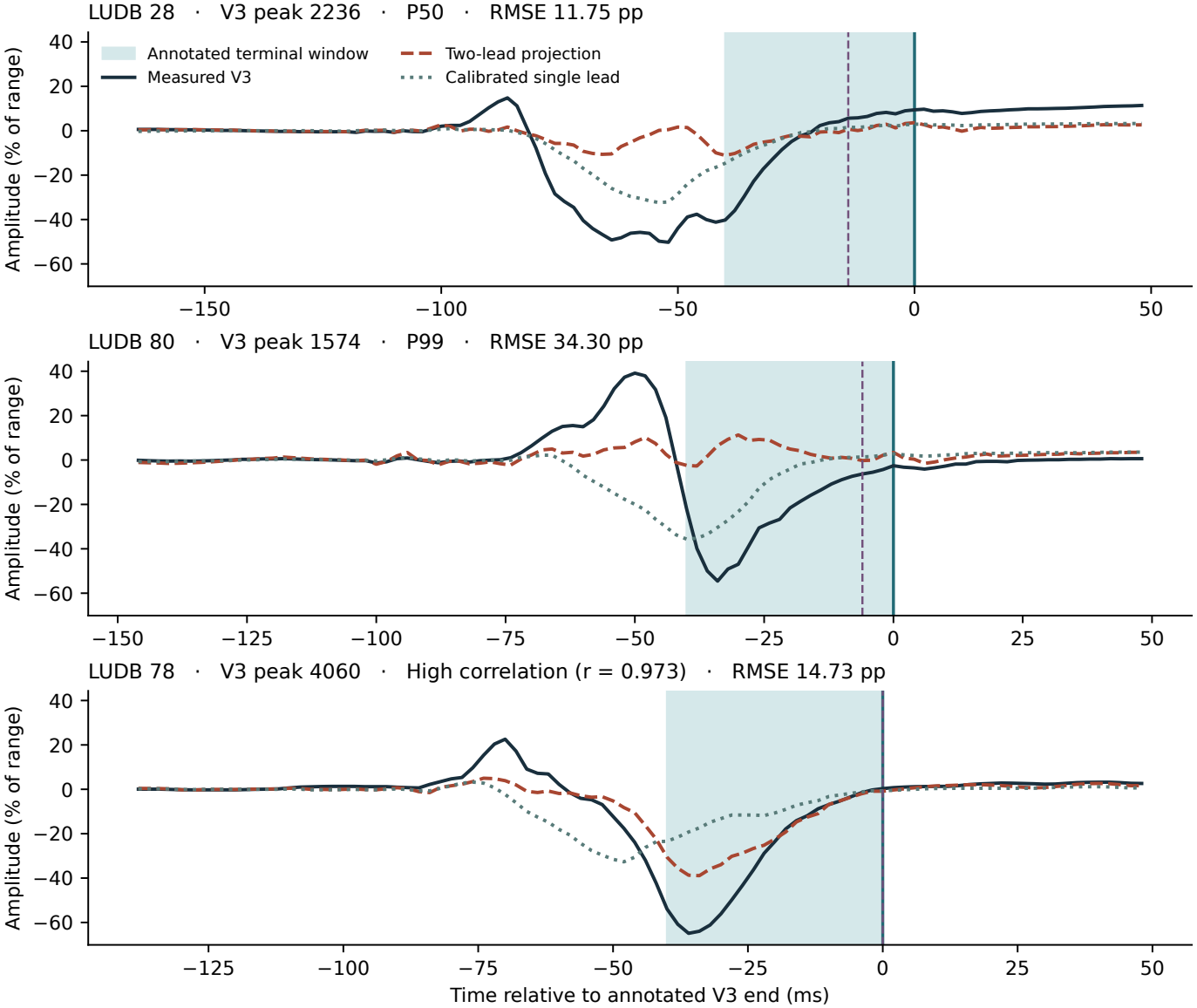
Measured and reconstructed V3 complexes. The first two panels were selected by proximity to the median and 99th percentile of V3 projection error. The third panel shows record 78, V3 peak 4060 (correlation 0.973, terminal RMSE 14.73 pp). Among V3 complexes with whole-QRS correlation of at least 0.95 and terminal RMSE of at least 10 pp, its error was tied for closest to the median. The lower record identifier determined the selection. Amplitudes are percentages of each channel’s 10-second range. Shading marks the 40 ms ending at the annotated V3 boundary. Time zero is the exclusive annotated stop. The dashed line marks the exclusive automatic stop and coincides with the annotated stop in the third panel. The single-lead fit had lower error than the projection in the first two examples.

## Discussion

V3 and V4 gained the most from combining the two precordial signals. Relative to the fitted single lead, mean terminal RMSE fell by 20.2% in V3 and 19.7% in V4 (Table 1). V5 also improved, although its single-lead estimate already had the lowest mean error. In V2 the fitted projection lay close to the V1 axis, and the single-lead comparator selected V1. The small observed difference in V2 is compatible with either a modest gain or a modest loss.

### What a fixed projection adds

The 3D ECG gives the angle a practical interpretation. A single trajectory of V1 and V6 over time yields a separate estimate for each intermediate lead by applying its preset angle at each sample. For example, the single-lead fit for V3 used V1, whereas the V3 projection assigned appreciable weight to both V1 and V6 (Section S2). We could therefore estimate relative terminal V3 amplitude in a new patient without using that patient’s V3 to choose the weights. The simultaneous V3 recording provided the reference against which to judge the estimate. Time orders the samples along the trajectory. Rotating the plotted figure leaves the numerical predictions unchanged (Section S7). Two unrestricted gains lowered overall mean RMSE by a further 0.23 pp. The lower bootstrap limit was close to zero and varied with the number of resamples (Sections S6 and S11). The study evaluated the numerical projections. Whether viewing the trajectory in three dimensions improves a reader’s interpretation remains an open question.

### Implications for reduced-lead recordings

If a recording retains V1 and V6 and the intermediate precordial leads are unavailable, the V3 and V4 projections offer the largest reduction in mean terminal error compared with the single lead selected in development. For V5, the improvement is smaller. The proposed use is to estimate the course of relative amplitude through the end of the QRS. A different question arises when interpretation depends on whether an individual beat contains a notch or slur [2]. Even with whole-QRS correlation of at least 0.95, terminal error reached one tenth of the lead’s recording range in 70 of 159 V3 pairs, 148 of 401 V4 pairs and 112 of 596 V5 pairs. These errors can reflect amplitude or timing differences. The threshold has no established diagnostic meaning. A blinded comparison of readers examining measured and reconstructed leads would be needed to establish whether those features can be recognized reliably. The distinction between the end of a complex in a single lead and its end across multiple leads has long been recognized in ECG delineation [11]. The choice of endpoint also changes which samples are judged. On the same 702 beats, projection RMSE was 13.02 pp with the input-defined end and 10.86 pp with the annotated target end. In Figure S1 a long run of identical digital samples complicates interpretation. Figure S2 shows a second V3 excursion outside the automatic window without a repeated 12-lead sample in that QRS. Both observations concern the recorded waveform. Neither establishes the physiological origin of the deflection. V3 had the largest estimated change in relative model benefit (Holm-adjusted *p* = 0.060).

Across test patients, mean RMSE fell from 10.64 to 3.33 pp when both coefficients could be fitted afresh to each measured target window (a reduction of 68.74%). The fit had access to the same 20 samples used to score it and could absorb noise as well as signal. Baseline uncertainty and similar V1 and V6 waveforms can both affect the residual. Its magnitude alone cannot identify an unmeasured electrical source.

## Limitations

This was an internal test in a single database already used by the investigators. Development and test patients were separate, but acquisition and annotation practices were shared. The normalization preserves relative amplitude within a channel and removes between-lead and between-patient differences in peak-to-peak amplitude. Recovering a missing target in clinical units would require its scale, which cannot be obtained from these data. Filtering in both directions and centred moving averages used later samples. Normalization and detector thresholds used the full ten-second record. The paired timing comparison further required a suitable automatic anchor and disproportionately lost beats from recordings labelled with ventricular pacing. Each lead has one published set of boundaries, precluding an estimate of inter-reader variability. The six annotations identify a common QRS by the stated rule. They do not establish an exact physiological end. Actual electrode loss, patient-level diagnostic decisions and effects on management were not studied.

## Conclusions

One preset angle per lead was sufficient to improve the offline estimate of relative amplitude in the last 40 ms of V3, V4 and V5 in held-out LUDB patients. The largest gains were in V3 and V4, where the V1/V6 projection reduced mean error by about one fifth compared with the fitted single lead.

The result for V2 remained uncertain. This gives a reduced-lead recording a way to estimate the terminal waveform of an unavailable precordial lead without fitting the projection to that patient. The voltage of an individual deflection and the diagnostic accuracy of a reconstructed notch or slur still require validation against directly measured leads.

## Supporting information

Supplementary appendix, analysis code and reproducibility data

## Data Availability

The ECG recordings and per-lead annotations are publicly available from the Lobachevsky University Electrocardiography Database (LUDB), version 1.0.1, on PhysioNet. The supplementary archive accompanying this manuscript contains the analysis code, fixed model coefficients, record assignments, and individual measurements used to reproduce the results.

## Glossary

*Terminal window*: the 20 samples scored at the end of a target QRS.
*Fixed projection*: the weighted sum of V1 and V6 at each sample, using one population angle for each target.
*Exclusive stop*: the first sample outside the scoring window.
*RMSE*: the square root of the mean squared normalized amplitude difference within one window.

## Data availability and source attribution

The ECGs and per-lead annotations are publicly accessible in LUDB version 1.0.1 on PhysioNet [8, 12, 13]. This article and its accompanying tables and figures contain information from the Lobachevsky University Electrocardiography Database, made available under the Open Data Commons Attribution License v1.0. The supplementary appendix gives the cohort flow, sensitivity analyses and annotated ECG example. The supplementary archive contains the Python programs, fixed model coefficients, record assignments and individual measurements. Its README.md explains how to reproduce the numerical results, figures and supplementary appendix from the public release.

## Acknowledgements

None.

