## Supplementary appendix, analysis code and reproducibility data for "Reconstructing the range-normalized terminal QRS from V1 and V6 using the time-embedded 3D ECG": Supplementary_Appendix.pdf

#### S1. Population and inclusion

**Table S1.** Each participant contributed one ECG. The released per-lead annotations are the joint reading of the two LUDB cardiologists. There is one set of boundaries per lead. V3 QRS markers first had to have a complete onset–peak–closing triplet. Each such triplet supplied one candidate. A surviving candidate contributes four target windows. The test and development groups are disjoint.

| | Development | Test | Total |
|---|---|---|---|
| Assigned patients | 120 | 80 | 200 |
| Evaluable patients | 118 | 80 | 198 |
| Marked V3 peaks | 1,088 | 743 | 1,831 |
| Incomplete V3 triplet | 9 | 5 | 14 |
| Complete V3 triplets | 1,079 | 738 | 1,817 |
| No six-lead match | 12 | 5 | 17 |
| Eligible beats | 1,067 | 733 | 1,800 |
| Target windows | 4,268 | 2,932 | 7,200 |

We ranked records by SHA-256. The UTF-8 input was `LUDB-QRS-confirmed-v1|` followed by the decimal record number. The first 120 were assigned to development and the remainder to testing. Two development records (7 and 34) had V3 annotations but no beat satisfying the six-lead rule. There was no replacement. All marked V3 peak symbols in this release were N; the symbol alone does not determine the clinical rhythm. The software exports the assignment for all 200 participants and the fitted coefficients.

Starting with all marked V3 peaks, 20/77 in pacing-labelled recordings and 11/1,754 in the other recordings were lost. Considering only peaks with a complete V3 triplet, 11 of 68 V3 beats in 9 records labelled with ventricular pacing were excluded, compared with 6 of 1,749 V3 beats in the other 191 records. The records wholly excluded were 7 and 34. These labels describe the recording; they do not adjudicate pacing in individual beats.

An earlier calibration study of time-embedded descriptors used the same 200 LUDB recordings. This overlap concerns the source cohort. The present development/test split and terminal-window analysis are specified separately in the accompanying protocol. See Bermejo Valdés AJ, *Calibration and processing sensitivity of time-embedded three-dimensional electrocardiographic descriptors: a two-dataset benchmark*, medRxiv preprint (2026), doi:10.64898/2026.09.14.26363023.

**Annotation and calibration.** For every accepted QRS symbol (N, V, A, /, L or R), the preceding and following annotation symbols had to be an opening and a closing boundary. For each V3 peak we searched each other precordial lead within 50 samples on either side. Both forward and reverse associations had to be unique. Onset, peak and closing annotations had to be ordered; all six QRS intervals had to intersect. The V3 anchor need not be the fiducial peak of the other leads. No terminal interval could start before that target's annotated onset. The overlap establishes beat identity but does not require equal lead-specific ends.

We decoded format-16 samples with the header baseline and gain. Reported amplitudes are percentages of the 10-second range of the corresponding lead (Section S10). One percentage point (pp) equals 10 nominal microvolts in these files. From the first baseline sample to the last precordial QRS stop, nonfinite values, missing code −32768 and rail 32767 caused rejection. Each lead also required a QRS excursion above 0.1 pp. Clipping below the rails could escape detection. For earlier V1/V6 onset $o$, baseline samples were $o-25, \ldots, o-6$. We verified the primary models' test-window errors with a second read of the signals using WFDB header scaling. Indices are zero-based and stops exclusive.

#### S2. Model restrictions and sensitivity analyses

**Population coefficients.** Columns contain the V1 and V6 coefficients for unit projection A, calibrated single lead B and unrestricted two-lead model C. All use squared-error training, equal patient weights and no intercept. A's angle is measured from the V1 axis.

| Target | Angle (°) | $A_1$ | $A_6$ | $B_1$ | $B_6$ | $C_1$ | $C_6$ |
|---|---|---|---|---|---|---|---|
| V2 | 18.43 | 0.9487 | 0.3161 | 0.7854 | 0.0000 | 0.9657 | 0.3338 |
| V3 | 40.82 | 0.7568 | 0.6536 | 0.4497 | 0.0000 | 0.8595 | 0.7868 |
| V4 | 67.83 | 0.3774 | 0.9260 | 0.0000 | 0.6855 | 0.5149 | 1.1227 |
| V5 | 79.09 | 0.1893 | 0.9819 | 0.0000 | 0.8617 | 0.2561 | 1.0842 |

For A, we evaluated 720 equally spaced angles and refined each local minimum before choosing the global minimum. B retained whichever of V1 or V6 gave the smaller loss after fitting its signed gain. C solved the two-variable normal equations. Model U chose the best of ±V1 and ±V6. For aligned-loss sensitivity all coefficients were estimated again on the same development patients, minimizing patient-weighted mean window RMSE. The coefficients are available in `model.json`. The primary A–B comparison allows different gain freedom; U–A compares unit gains and B–C compares free gains.

**Table S2.** Differences in RMSE (pp) on identical confirmed test windows, with patient-bootstrap 95% intervals. Positive values favour two inputs. The last two columns use training loss aligned with the evaluation endpoint.

| Target | U minus A | B minus C | B minus A, aligned | B minus C, aligned |
|---|---|---|---|---|
| Overall | 2.06 [1.44, 2.68] | 2.17 [1.41, 2.90] | 1.75 [1.27, 2.21] | 1.99 [1.37, 2.60] |
| V2 | -0.09 [-1.19, 1.05] | 0.37 [-0.36, 1.09] | 0.34 [-0.12, 0.81] | 0.37 [-0.13, 0.87] |
| V3 | 3.81 [1.59, 6.21] | 3.38 [2.09, 4.62] | 2.77 [1.58, 3.97] | 2.90 [1.53, 4.27] |
| V4 | 3.17 [1.46, 4.71] | 3.27 [2.08, 4.39] | 2.67 [1.57, 3.70] | 3.11 [1.92, 4.23] |
| V5 | 1.36 [0.44, 2.24] | 1.67 [1.00, 2.30] | 1.22 [0.66, 1.74] | 1.58 [0.95, 2.18] |

**Table S3.** Fixed-model primary reduction, sensitivity to excluding the one test participant above the 99th percentile of mean A error, and the out-of-fold reduction across all 198 eligible participants. Units are pp. For five-fold validation, subjects at sorted ranks $i$ were assigned fold $i$ mod 5, starting from zero. Each held-out fold was predicted with coefficients estimated on the other four folds. The out-of-fold rows summarize a secondary point estimate.

| Target | Main test | Trimmed test [95% CI] | Out of fold |
|---|---|---|---|
| Overall | 1.94 | 1.96 [1.34, 2.58] | 2.00 |
| V2 | 0.35 | 0.30 [-0.37, 0.98] | 0.56 |
| V3 | 3.23 | 3.17 [2.06, 4.26] | 3.34 |
| V4 | 2.85 | 2.97 [1.94, 3.97] | 2.85 |
| V5 | 1.35 | 1.41 [0.83, 1.99] | 1.25 |

#### S3. End-of-QRS sensitivity

Only beats with six cardiologist annotations entered the paired comparison. We ran the detector on V1 and V6 over the entire ten-second record at 500 Hz. Both bandpasses used third-order Butterworth designs in second-order sections. We applied each filter forwards and backwards along time with `sosfiltfilt`, odd endpoint extension and the SciPy default padding length (21 samples per end here). For the 8–30 Hz output we averaged the squared values across channels, took a centred 13-sample moving mean and then the square root. Both moving means in this detector used `uniform_filter1d` with `mode="reflect"` and zero origin. For the resulting energy envelope, let $m$ be its median over the record and $q$ its 99th percentile minus $m$. We used `find_peaks` with height at least $m + 0.17q$, prominence at least $\max(0.3\text{ pp}, 0.14q)$ and minimum separation of 125 samples. The code uses nominal header units (10 per pp).

We first-differenced each 1–45 Hz filtered input along time, prepending zero to that filtered channel. We averaged the squared differences across inputs, took a centred five-sample moving mean and then its square root. For peak $p$, the quiet threshold was the largest of 0.1 pp per sample, 2.4 times the record-wide 22nd percentile of this activity and 0.085 times its maximum from $p - 18$ through $p + 18$. After peak detection we excluded $p < 95$ or $p + 61 \geq N$ for a record of $N$ samples. On each side we searched outwards at distances 8 through 55. We retained the first index $a$ with six successive values at or below threshold ($a - 5$ through $a$ on the left, $a$ through $a + 5$ on the right). Both boundaries were required, with a separation of 20–110 samples and a right boundary of at least 30. The right boundary was the exclusive stop. The original unfiltered amplitudes supplied the reconstruction scores.

In development and test, respectively, the detector found 1,340 and 906 peaks, of which 30 and 28 lay at excluded edges. After the edge rule, 1,310 and 878 remained. Of these, 1,276 and 833 had valid boundaries and passed the signal checks in both inputs and all four measured targets. The sole eligible anchor had to lie inside the intersection of all six annotated QRS intervals for a beat. This yielded 1,043 development beats in 118 participants and 702 test beats in 77 participants. Of the 131 quality-eligible test detections without a match, 124 fell outside the span of the annotated V3 peaks in their record. The available annotations do not classify all unmatched detections as false positives.

**Table S4.** Identical 702 held-out beats in 77 patients. Bias is input minus target offset. Changes in each model's RMSE are annotated minus automatic. The marginal 95% percentile intervals were obtained by resampling patients.

| Target | Bias (ms) | Mean absolute (ms) | Change A [95% CI], pp | Change B [95% CI], pp |
|---|---|---|---|---|
| Overall | -4.31 | 8.10 | -2.16 [-2.98, -1.42] | -2.45 [-3.31, -1.63] |
| V2 | -5.02 | 8.98 | -1.73 [-2.73, -0.76] | -1.33 [-2.26, -0.41] |
| V3 | -7.17 | 8.81 | -3.86 [-5.17, -2.71] | -4.93 [-6.49, -3.48] |
| V4 | -4.45 | 7.69 | -2.55 [-3.90, -1.31] | -2.99 [-4.52, -1.58] |
| V5 | -0.62 | 6.94 | -0.48 [-1.22, 0.17] | -0.54 [-1.29, 0.12] |

**Endpoint convention.** For target closing annotation $k$ the exclusive stop is $k + 1$. For the detector it is the first qualifying quiet sample $f$. The main comparison uses samples $k - 19, \ldots, k$ versus $f - 20, \ldots, f - 1$. The reported offset is $2[f - (k + 1)]$ ms. Section S9 varies the automatic stop by one sample.

#### S4. Absolute error and reproducibility

The retrospective within-window fit D solved least squares on the measured target and the 20-by-2 matrix of input samples at that same time. Mean RMSE was 10.64 pp for A, 10.41 pp for C and 3.33 pp for D. The ratio of patient-weighted mean squared D residual to mean squared A error was 11.75%. The median across test patients of the within-patient median condition number of the input matrix (beat–lead windows) was 5.83. All 2,932 test matrices had rank two. In 34 windows the spectral condition number exceeded 30 (maximum 57.92). Within each window we computed the Pearson correlation between the first and last 19 D residuals, shifted by one sample. The median over 2,932 windows was 0.76. Adding an intercept to the within-window fit gave a patient-weighted mean RMSE of 2.37 pp. Fitting each contiguous 10-sample half and testing on the opposite half gave 17.43 pp using all 20 out-of-fit predictions per window. Nearby samples carry correlated errors. Similar inputs can make coefficient estimates unstable, and fitting to the target can absorb noise, so D is a descriptive lower bound for this functional form.

**Table S5.** Coexistence of high Pearson correlation across the target's entire annotated QRS and high projection RMSE in its terminal 40 ms. The joint denominator is all 2,932 test beat–lead pairs. The conditional denominator is the number meeting the correlation threshold. Each observation has equal weight in this table.

| Min. $r$ | Min. RMSE (pp) | High $r$ | Both | Joint (%) | Conditional (%) |
|---:|---:|---:|---:|---:|---:|
| 0.90 | 5.00 | 1,940 | 1,324 | 45.16 | 68.25 |
| 0.90 | 10.00 | 1,940 | 643 | 21.93 | 33.14 |
| 0.90 | 20.00 | 1,940 | 105 | 3.58 | 5.41 |
| 0.95 | 5.00 | 1,498 | 1,000 | 34.11 | 66.76 |
| 0.95 | 10.00 | 1,498 | 432 | 14.73 | 28.84 |
| 0.95 | 20.00 | 1,498 | 73 | 2.49 | 4.87 |
| 0.99 | 5.00 | 571 | 308 | 10.50 | 53.94 |
| 0.99 | 10.00 | 571 | 123 | 4.20 | 21.54 |
| 0.99 | 20.00 | 571 | 22 | 0.75 | 3.85 |

**Reproduction.** Unzip the supplementary archive and follow `README.md`. The analysis reads the public LUDB recordings and annotations, checks their hashes, and writes the beat windows, model coefficients and patient-level results. A second read verifies both models' errors on the annotated and automatic windows. The same workflow generates the figures and this appendix. The file index identifies the inputs and outputs used for each analysis. The first two traces in Figure 2 are nearest the median and 99th percentile of V3 projection error. Figure 1 uses the median case. Distances within $10^{-11}$ pp of the minimum are tied and ordered by ascending patient identifier, then V3 peak sample. In the 70 V3 pairs meeting both thresholds, this rule selects record 78, peak 4060 for Figure 2C.

To locate a window in a source ECG, select a row of `beat_windows.csv`. For target V3 take the measured voltages from `V3_stop` minus 20 up to, but excluding, `V3_stop`. Read V1 and V6 at precisely those indices. Centre each channel with its own median over the stored baseline interval before applying the two coefficients. Match the result to `test_observations.csv` by patient, reference peak and target.

#### S5. Visual check of QRS identity and timing

**Figure S1.** Six simultaneously measured precordial leads from held-out LUDB record 99. The annotated V3 peak is sample 2088. Each panel shows the cardiologist's marks in that lead and the common end estimated from V1 and V6. The solid end line marks the first sample after that lead's closing annotation. The dashed line marks the common automatic stop. The shaded interval comprises the preceding 20 measured samples.

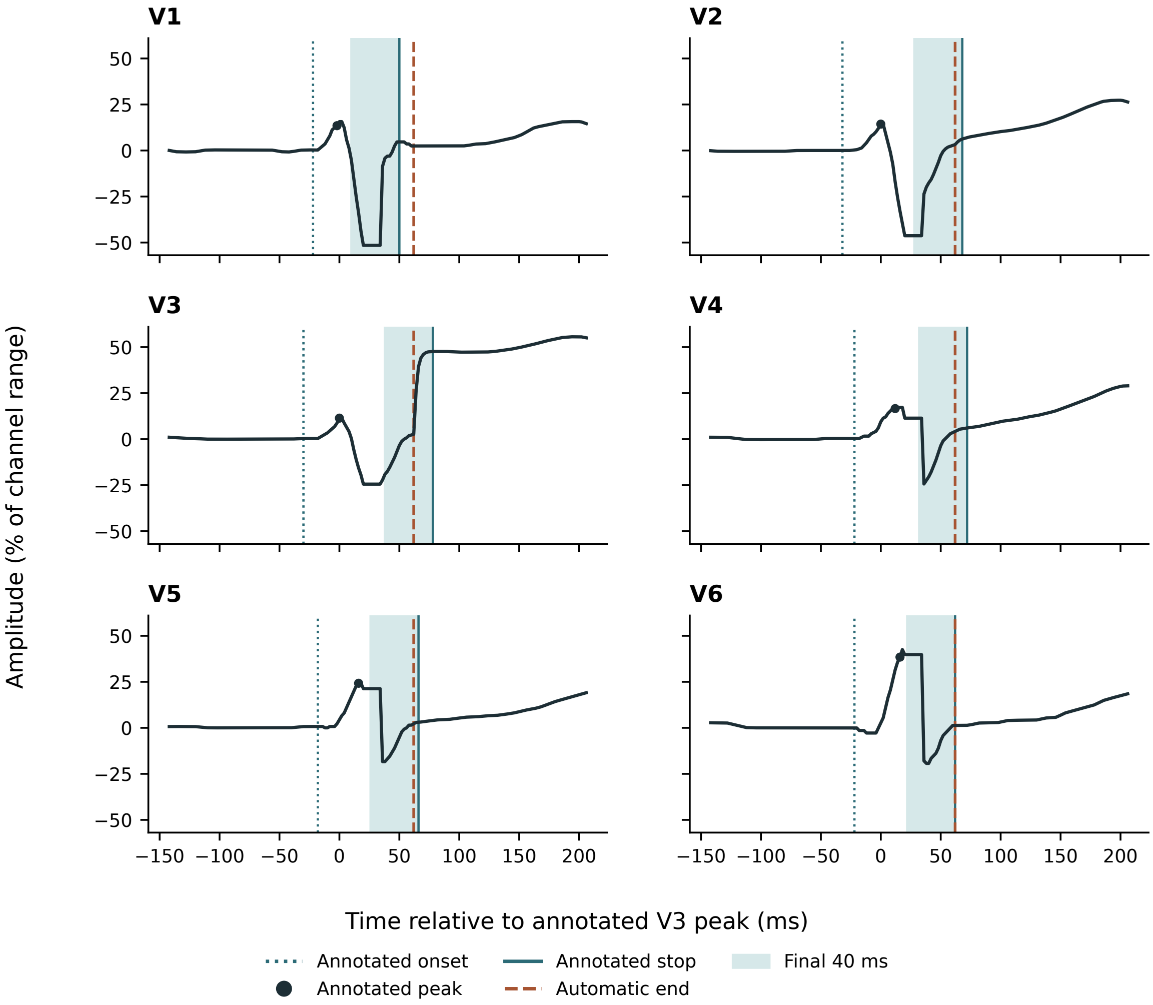


In V3 the automatic window ended 16 ms before the annotated end. The measured peak-to-peak excursion in the omitted part of the annotated terminal window was 44.92 pp. This example was chosen for its timing failure. Among paired test beats with an automatic end at least 10 ms before the V3 end, we selected the largest V3 excursion in the omitted part of the final 40 ms, breaking ties by record number and peak sample. From samples 2098 to 2105 inclusive, the digital vector across all 12 leads is identical. An acquisition or processing artefact is plausible; its cause is unknown. This is an excursion in the recorded signal. The figure establishes neither the physiological origin of that excursion nor the prevalence of clinical notching.

#### S6. Paired inference and uncertainty in fitting

For each patient $i$ and lead $j$ we averaged the beat-level RMSE difference $d_{ij} = \overline{\mathrm{RMSE}_B - \mathrm{RMSE}_A}$. For the primary comparison $d_i = \sum_{j=2}^{5} d_{ij}/4$. We tested $H_0 : E(d_i) = 0$ with $t = \bar{d}/(s_d/\sqrt{n})$ and $n-1$ degrees of freedom. Inference assumes independent patients and a sufficiently regular distribution of the sample mean. The same calculation by lead used Holm correction for four two-sided tests. We added these tests after examining the original descriptive results.

**Table S6.** Fixed-model tests in 80 patients. Simultaneous intervals use the Bonferroni $t$ critical value ($1 - 0.05/8$ quantile, 79 degrees of freedom). The last column gives marginal percentile intervals from a separate bootstrap with refitting. Intervals are in pp.

| Target | df | $t$ | Raw $p$ | Holm $p$ | Simultaneous CI | Refit CI |
|---|---|---|---|---|---|---|
| Overall | 79 | 6.04 | $4.82 \times 10^{-8}$ | – | – | [1.29, 2.63] |
| V2 | 79 | 0.98 | 0.330 | 0.330 | [-0.56, 1.27] | [-0.45, 1.14] |
| V3 | 79 | 5.71 | $1.87 \times 10^{-7}$ | $7.49 \times 10^{-7}$ | [1.78, 4.67] | [2.04, 4.54] |
| V4 | 79 | 5.46 | $5.23 \times 10^{-7}$ | $1.57 \times 10^{-6}$ | [1.52, 4.18] | [1.80, 3.87] |
| V5 | 79 | 4.58 | $1.73 \times 10^{-5}$ | $3.46 \times 10^{-5}$ | [0.60, 2.11] | [0.74, 1.94] |

In 4,000 replicates we sampled 118 development patients and 80 test patients independently, with replacement within each group. Each draw retained all the selected patients' beats. We re-estimated the projection angle and re-selected the single lead and its gain using only the resampled development patients. We scored these fits in the resampled test patients. The global 95% interval was [1.29, 2.63] pp. For the original fixed models, the range of overall estimates when omitting each test patient in turn was [1.87, 2.06] pp. Excluding all three test recordings carrying a ventricular pacing label gave 1.89 [1.24, 2.52] pp in the other 77 patients. Of 16 eligible test beats in records labelled with ventricular pacing, only five, all from one record, had a matched automatic end. Performance specific to paced beats remains uncertain. After removing the entire recording selected for its large terminal excursion in Figure S1, the mean benefit was 1.93 [1.29, 2.54] pp (79 patients).

**Table S7.** Change in advantage on replacing the automatic end with the annotated end. The global test and four secondary lead tests used 77 patients (76 degrees of freedom); Holm adjustment was confined to the four leads. The displayed $t$ intervals are marginal.

| Target | $t$ | Raw $p$ | Holm $p$ | 95% CI (pp) |
|---|---|---|---|---|
| Overall | -1.87 | 0.065 | – | [-0.60, 0.02] |
| V2 | 1.68 | 0.098 | 0.293 | [-0.08, 0.89] |
| V3 | -2.49 | 0.015 | 0.060 | [-1.93, -0.21] |
| V4 | -1.65 | 0.103 | 0.293 | [-0.97, 0.09] |
| V5 | -0.60 | 0.552 | 0.552 | [-0.28, 0.15] |

The exploratory A-minus-C mean difference was 0.23 pp. For this contrast the lower percentile limit lay close to zero and crossed it for a different 5,000-sample draw (Section S11). The analyses of gain restrictions, alternate fitting loss and influence are sensitivity estimates. Timing and refitting were evaluated on the same source cohort. Neither constitutes external replication.

**Earlier detector-defined sample.** Applying the earlier lead-by-lead rule produced 7,031 beat–lead observations. In 7,027 the input anchor fell inside the target QRS. All four exceptions arose at sample 3608 in record 74. Among the earlier observations we could match to both annotated input onsets in the current test patients, the fixed models gave a reduction of 1.92 [1.27, 2.53] pp (2,808 observations in 77 patients). The 2,808 observations reuse the same participants, leads and acquisition times as the matched timing analysis. This historical comparison supplies no independent replication.

#### S7. Physical meaning of the projection

Let $x_1(t)$ and $x_6(t)$ be the two baseline-corrected, range-normalized amplitudes in pp. The curve $\gamma(t) = (x_1(t), x_6(t), t)$ has two normalized amplitude coordinates and a time coordinate measured in milliseconds. At each $t$, the predicted normalized amplitude is $\widehat{v}_j(t) = (\cos\theta_j, \sin\theta_j) \cdot (x_1(t), x_6(t))$. The coefficients are dimensionless and the dot product is in pp. Time indexes the pair. It carries no fitted voltage coefficient. Changing its display units cannot change this prediction.

Figure 1 of the main article illustrates this construction in LUDB record 28 (V3 peak at sample 2236). The file `trajectory_3d.csv` contains the two normalized inputs in nominal header units, time, the measured V3 and the projected V3 at each displayed sample. The verification script compares the projected amplitudes with the values obtained independently from the source recording.

For a change of input scale $x' = Sx$, unrestricted weights $w' = S^{-\top}w$ preserve the prediction. These weights need not have unit norm. As a numerical sensitivity check, doubling V1 alone and refitting in development changed the B-minus-A advantage to 1.48 pp overall and -1.80 pp in V2. The target scale was held fixed. The displayed angle therefore applies to the stated channel normalization. A three-component dipole model would supply at most two independent instantaneous equations from V1 and V6, so those two measurements alone cannot identify its three spatial components. No dipole inversion, anatomical rotation or measured orthogonal vectorcardiogram was used. The measured target electrode tests the prediction of relative terminal amplitude. Recognition of a notch or slur would require a separately adjudicated clinical endpoint.

#### S8. Residual error in highly correlated reconstructions

**Table S8.** Test beat–lead pairs with full-QRS Pearson $r \geq 0.95$ and projection RMSE $\geq 10$ pp over the annotated terminal window. The joint percentage uses all pairs in the row. The conditional percentage uses only pairs meeting the correlation threshold. The marginal 95% CIs for the conditional percentages come from 5,000 resamples of whole test patients, recalculating the ratio of counts in each sample. These are beat–lead frequencies. 75 of 80 test patients contributed at least one high-correlation pair. The 159 high-correlation V3 pairs came from 27 participants. Globally, 28.84% of high-correlation pairs crossed the error threshold, compared with 60.32% of pairs below the correlation threshold. Thus correlation was informative in aggregate but did not guarantee low terminal error in a given pair.

| Lead | All pairs | High $r$ | Both | Joint (%) | Conditional (%) | 95% CI (%) |
|---|---|---|---|---|---|---|
| Overall | 2,932 | 1,498 | 432 | 14.73 | 28.84 | [22.65, 35.44] |
| V2 | 733 | 342 | 102 | 13.92 | 29.82 | [18.77, 42.50] |
| V3 | 733 | 159 | 70 | 9.55 | 44.03 | [28.12, 62.31] |
| V4 | 733 | 401 | 148 | 20.19 | 36.91 | [25.54, 48.83] |
| V5 | 733 | 596 | 112 | 15.28 | 18.79 | [11.28, 27.23] |

The RMSE averages amplitude discrepancies over the entire 40-ms window. It does not count missed notches or measure J-point displacement in individual beats.

### S9. One-sample endpoint sensitivity

We moved the exclusive automatic stop one sample earlier or later, preserving the 702 beats, 77 participants, measured signals, baseline and frozen coefficients of the matched analysis. This additional descriptive calculation tests sensitivity to how the first low-slope sample is treated. It uses the same data as Table S4.

**Table S9.** Single-lead error minus projection error (pp). Positive values favour the projection. The column headings indicate the displacement from the original automatic stop. The software supplies each model's absolute RMSE at all three positions.

| Target | −2 ms | Original stop | +2 ms |
|---|---|---|---|
| Overall | 2.33 | 2.22 | 2.10 |
| V2 | -0.04 | -0.07 | -0.08 |
| V3 | 4.55 | 4.17 | 3.80 |
| V4 | 3.38 | 3.34 | 3.28 |
| V5 | 1.41 | 1.42 | 1.40 |

The point estimate kept its sign across the three positions in each lead. V3 showed the largest absolute change in the estimated advantage. These summaries depend on the same annotated cohort and apply to the scoring convention used here.

### S10. Scaling, signal quality and timing examples

For digital counts $d_j(t)$, header baseline $b_j$ and gain $g_j$, the stored nominal amplitude is $(d_j(t) - b_j)/g_j$ mV. We computed the full-record digital range independently for every channel. In 2,400 of 2,400 channels $g_j = \max_t d_j(t) - \min_t d_j(t)$. After centring, the plotted signal is $100(d_j(t) - \text{median}_{\text{baseline}} d_j)/g_j$ percent of that channel's range. Thus the unit-norm angle applies to separately normalized inputs. The figures and article express differences of normalized amplitudes in percentage points. The numerical files from the original analysis retain their nominal units so that every conversion can be traced. Neither the public headers nor the distribution notes provide an independently verified common scale for patient voltages.

As a cross-check, the time variation in the digital residual II−I−III was zero in 200/200 recordings. On applying the separate channel gains, the median within-record standard deviation of that residual was 60.09 nominal microvolts. This observation illustrates how the header scaling changes cross-lead amplitudes, without determining the instrument's native voltage calibration.

We counted consecutive identical digital vectors across all 12 leads within the union of the six annotated precordial QRS intervals. Fourteen of 733 test beats had a run of at least five samples. Excluding those beats (719 beats in 80 patients), the frozen-model advantage was 1.96 pp (95% patient-bootstrap CI 1.32 to 2.56). The record in Figure S1 contained the only test beat with a run of at least eight samples. Repeated values need not imply a specific acquisition fault.

**Figure S2.** Additional timing example without a repeated digital vector. Of the paired test beats with an automatic stop at least 10 ms early in V3, we excluded beats with a run of five or more identical 12-lead samples and selected the largest remaining V3 excursion after the automatic stop within the final annotated 40 ms. Ties were broken by record and peak index. In record 71 (V3 peak 4218), the automatic stop was 4244 and the V3 stop was 4259. Between them, V3 spanned 24.72 pp of its recording range. Both fitted predictions and the recorded V3 are shown. This example was selected for discrepancy and gives no estimate of prevalence.

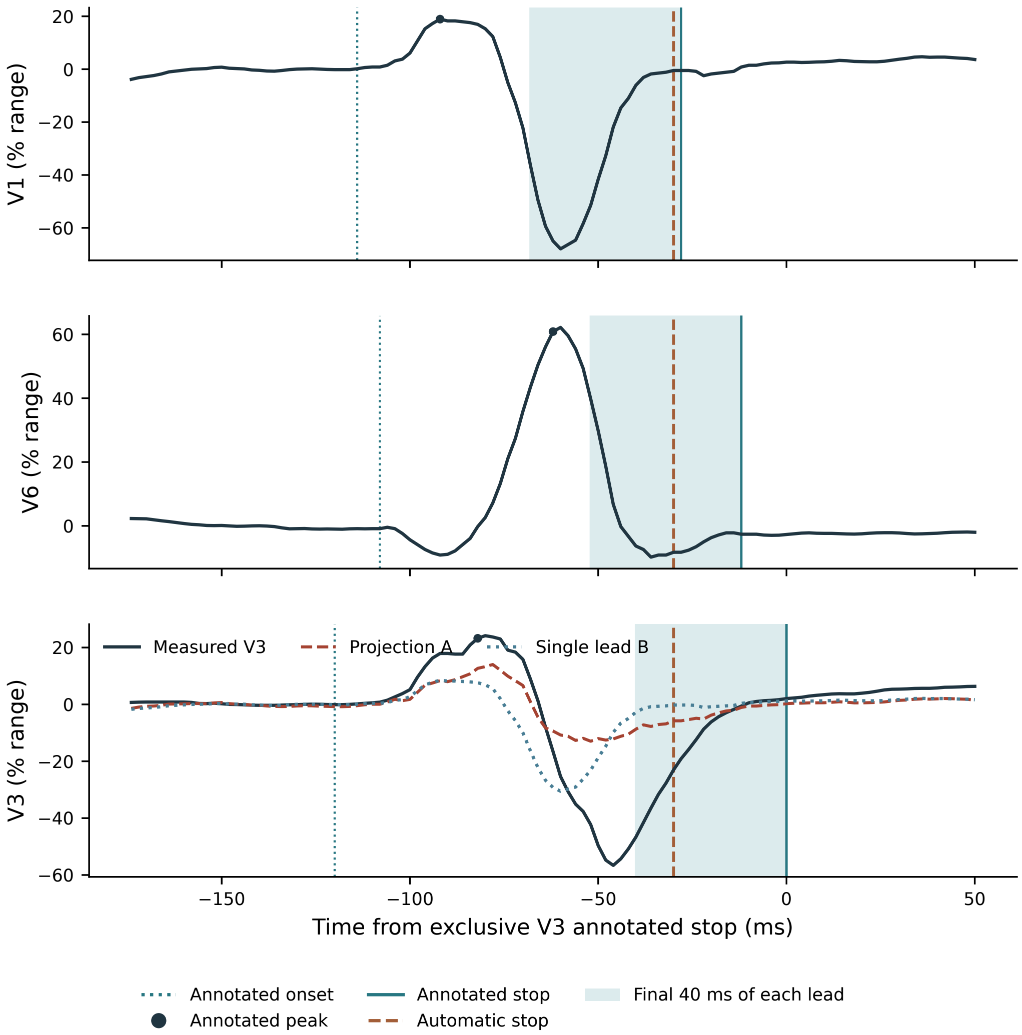


#### S11. Baseline, window placement and comparator sensitivity

The primary centring interval is a local statistical reference and can extend into an annotated QRS if a target starts before V1 and V6. This affected 155/1,067 development beats and 107/733 test beats. In V2 through V5 the test counts were 51, 60, 44 and 18, respectively. We retained the original terminal windows and examined two alternatives: the earliest of all six annotated onsets with the original 10-ms gap, and the earlier V1/V6 onset with a 30-ms gap. For each we scored the original fixed coefficients and repeated fitting only in development. The six-lead baseline requires the target annotations and serves as a sensitivity analysis. With the 30-ms input-only gap, four test beats still had baseline overlap.

**Table S10.** Mean B-minus-A difference (pp) on the original 80 test participants. Refit columns use coefficients estimated on the original development partition with the specified baseline.

| | Primary | Six, fixed | Six, refit | Inputs, fixed | Inputs, refit |
|---|---|---|---|---|---|
| Overall | 1.94 | 1.94 | 1.93 | 1.94 | 1.89 |
| V2 | 0.35 | 0.34 | 0.33 | 0.38 | 0.32 |
| V3 | 3.23 | 3.23 | 3.23 | 3.32 | 3.26 |
| V4 | 2.85 | 2.83 | 2.83 | 2.75 | 2.71 |
| V5 | 1.35 | 1.35 | 1.35 | 1.31 | 1.29 |

For the 702 matched beats, the automatic-stop analysis originally used the annotated input onset for baseline timing. We also used the automatic onset to centre only the inputs, keeping the scored target reference unchanged, and then to centre all channels. The global advantages were 2.22, 2.24 and 2.20 pp, respectively. These comparisons remain conditional on a known, matched QRS. Ending all four windows at the latest annotated precordial stop instead of the local target stop changed 2,229 of 2,932 windows and gave a global B-minus-A advantage of 1.66 pp in all 733 beats. Of the 2,932 latest-stop windows, 703 lay wholly inside the local QRS, 2,229 extended past its closing sample and 13 lay entirely after it. This sensitivity therefore scores a different segment from the target-specific terminal window. Of 31 eligible beats omitted from the timing comparison, 28 had an energy peak but no eligible pair of limits within the intersection, and three had no energy peak there.

**Table S11.** Position of the exclusive automatic stop relative to the annotated target stop. Counts are per beat. The last column counts pairs whose two 20-sample windows do not overlap.

| Target | Early | Coincident | Late | No overlap |
|---|---|---|---|---|
| V2 | 468 | 50 | 184 | 8 |
| V3 | 524 | 66 | 112 | 13 |
| V4 | 431 | 79 | 192 | 13 |
| V5 | 285 | 74 | 343 | 9 |

The centring interval overlapped an annotated P wave in at least one chest lead in 454/733 test beats. After excluding all annotated P and QRS samples within that interval, 665 beats from 80 patients retained at least five samples in every lead. With fixed coefficients, the B-minus-A advantage in those same beats was 1.954 pp with the original median and 1.945 pp with the masked median. For a separate comparator sensitivity we centred each candidate single input on its own marked onset, reselected the input and its gain in development, and held the target reference and A fixed. The mean advantage was 1.99 pp across all test patients. The comparator still assumed that the selected lead's onset was known.

For the exploratory contrast of unit-norm A versus unrestricted two-input C, the estimated mean reduction in error was 0.23 pp. With 200,000 patient resamples, the percentile interval was [0.003, 0.441] pp. Its lower limit was close to zero and varied across shorter draws. The supplementary file `bootstrap_precision.csv` gives the original draw, an alternative draw of 5,000 resamples and a 200,000-replicate interval. These comparisons were added after inspection of the results.

#### Source attribution

The recordings and annotations were obtained from Lobachevsky University Electrocardiography Database, version 1.0.1, by Kalyakulina and colleagues (PhysioNet, 2021; doi:10.13026/eegm-h675). They are available under the Open Data Commons Attribution License v1.0. The ECG examples and all derived tables use that release.
